# Disruption of outdoor activities caused by wildfire smoke shapes circulation of respiratory pathogens

**DOI:** 10.1101/2023.07.09.23292078

**Authors:** Beatriz Arregui García, Claudio Ascione, Arianna Pera, Davide Stocco, Boxuan Wang, Eugenio Valdano, Giulia Pullano

## Abstract

The consequences of wildfires on public health extend beyond injury. Smoke can traverse vast distances, compromising air quality in unaffected areas and exacerbating chronic respiratory conditions. But smoke may affect the circulation and burden of communicable diseases, too. The disruption in air quality and adherence to safety guidelines can impact the time people spend indoors, and this in turns may increase exposure to airborne pathogens like influenza, SARS-CoV-2, RSV. However, the quantification of such disruptions and their implications for the transmission of respiratory diseases remain unclear. Here we study the effects of smoke generated by severe wildfires in the U.S. states of California, Oregon, Washington in September 2020. We assess the impact on human behavior and the potential consequences for the emergence of respiratory diseases. Our findings reveal a significant shift towards indoor activities in counties within Oregon and Washington during wildfires. However, a discernible change in mobility patterns is not evident in California. This discrepancy may arise from the familiarity of Californian residents with wildfires and air quality index alerts, which have become integrated into their daily routines. Consequently, their mobility patterns may be less affected during such incidents compared to individuals in other regions. We then use a deterministic compartmental model of epidemic spread to quantify the impact of the describe behavioral changes on epidemic circulation. We found that counties with disrupted air exhibited higher cumulated and peak incidence of cases compared to unaffected counties, with the exception of California. Additionally, we found that flu-like epidemics – low reproduction ratio and short generation time – are most affected by the behavioral changes under study. Our findings may help improve public health response in a context of larger, more frequent wildfires triggered by climate change.

## 1 Introduction

Climate change is causing larger, longer and more devastating wildfires everywhere, and this is especially visible in the United States of America. Adopting this perspective, the New York Times defines wildfires ravaging the West Coast as climate fires [1]. The 2020 wildfire season in the Western United States occurred during the warmest period since global climate records began in 1880, and it was one of the seasons with the highest number of wildfires in the last two decades. Specifically, in August 2020, thunderstorms triggered multiple wildfires across the states of Oregon, Washington, and California which were then followed by additional fire outbreaks along the West Coast in early September [2].

Wildfires cause harm beyond the direct destruction they cause: atmospheric circulation carries the smoke to areas unaffected by the wildfires, compromising air quality over large swathes of land [3]. Smoke exacerbates pre-existing respiratory chronic diseases, increasing their morbidity and mortality [4]. For this reason, the Centers for Disease Control (CDC) provides guidelines to limit outdoor physical activity when and where wildfire smoke makes the air unsafe [5]. But wildfires may threaten human health in other ways, by changing behavioral patterns [6, 7]. Directly perceived air quality deterioration and guidelines induce changes in the ability and willingness of individuals to engage in outdoor activities, altering recurrent human behavior habits, and these behavioral changes may affect the spread of communicable disease [8–14]. This is especially true in the case of airborne respiratory pathogens: influenza, SARS-CoV-2, and Respiratory Syncytial Virus (RSV). Their spread is greatly driven by the social behavior of individuals [15], and indoor activities facilitate transmission and play a significant role in shaping the variability of disease dynamics [16]. While seasonal human behavior changes and their impact on hampering or mitigating epidemic activity have been largely studied [17–19], the impact of Wildfire smokes on human behaviors and the cascade effect for respiratory disease transmission is still unclear.

To address this question, we use a metric of the relative tendency of human interactions to take place indoors at a detailed spatial and temporal scale throughout the United States, as defined in the study published in [16]. We select the U.S. counties with the highest Air Quality Index during the 2020 Western United States wildfire season and quantify the impact of deteriorated air quality on seasonal indoor and outdoor activities. Subsequently, we incorporate this information into an infectious disease transmission model and compare the results to scenarios where indoor activity remains at baseline levels. Our research provides evidence of a shift in human behavior across states in the U.S. during wildfires, enhancing our understanding of the interplay between human behavior and the risk of infection in the context of climate change.

This report describes the results of an educational project carried out in 3 days’ time within the framework of the workshop Complexity72h. As such, it provides the starting point for additional research into a problem of great public health relevance, and which climate change is likely to further exacerbate.

## 2 Methodology

### 2.1 Data

To select affected counties by wildfire smoke, we analyze the daily Air Quality Index (AQI) in all US counties [20]. The AQI monitors the levels of significant air pollutants that are regulated under the Clean Air Act [21]. Data are collected by Environmental Protection Agency (EPA) and are available on their website[22].

AQI values are made available daily and can be split into the following intervals:

- 0 ≤ *AQI* ≤ 50. **Good**: air quality is satisfactory, and air pollution poses little or no risk.
- 50 < *AQI* ≤ 100. **Moderate**: air quality is acceptable. However, there may be a risk for some people, particularly those who are unusually sensitive to air pollution.
- 100 < *AQI* ≤ 150. **Unhealthy for sensitive groups**: members of sensitive groups may experience health effects. People are less likely to be affected.
- 150 < *AQI* ≤ 200. **Unhealthy**: some persons may experience health effects; members of sensitive groups may experience more serious health effects.
- 200 < *AQI* ≤ 300. **Very unhealthy**: health alert, the risk of health effects is increased for everyone.
- *AQI* > 300. **Hazardous**: health warning of emergency conditions: everyone is more likely to be affected.

In our analysis, we employ the weekly average AQI for each county in the continental United States of America. We focus only on the counties of the states of Oregon (OR), Washington (WA), and California (CA) since these three states were the most affected by smoke caused by more than 100 wildfires affecting the Western US states in September 2020 [23]. As the starting date of the phenomenon, we refer to 10 September 2020 in our analyses since it is the date on which the smoke started to spread over the West Coast. For the analyses focusing on variation over a broader timeframe, we consider the period between July 1, 2020 and November 1, 2020.

We select the counties in OR, WA and CA reporting an AQI greater than 150 (AQI level corresponding at least to “unhealthy”) for at least three days during the period between July 1, 2020 and November 1, 2020. We then filter such counties by selecting those that are part of the same indoor activity “Northern” cluster as defined in [16]. For each state, we consider the top 5 counties by population ^1^. Such a selection process results in the definition of the following affected counties:

- **OR**: Multnomah, Washington, Clackamas, Lane, Marion.
- **WA**: King, Spokane, Clark, Thurston, Yakima.
- **CA**: Plumas.

We then define as *unaffected* all U.S. counties characterized by a *good* or *moderate* AQI level during the studied time window. We select unaffected counties with population size in the top 25% of the distribution (i.e. population size *≥* 67976), for a total of 50 unaffected counties. Those counties will be considered as a baseline in our analyses.

To analyze human behavior, we consider the weekly indoor seasonality index *σ*_*it*_ by county computed in Ref. [16] and available here ^2^. The index takes values higher (lower) than 1 if, on average, indoor (outdoor) activities are preferred to outdoor (indoor) ones. The data is based on SafeGraph Weekly Patterns data [24] from 2018 to 2021, and 4.6 million POIs are sampled in all years of our study from all locations where people can spend their time, excluding home locations. The dataset is anonymized, and information on individual mobile devices is omitted. Data are representative across various demographics, including race/ethnicity, educational attainment, and income [25], and it has also been shown not to be significantly biased by age and gender [26], except for individuals younger than 16 years of age are not included [27].

### 2.2 Regression Discontinuity of indoor activity seasonality

To detect discontinuities in the seasonal pattern of indoor activity, we employ the Regression Discontinuity (RD) approach [28–31]. It is a statistical method used to estimate the causal effect of a treatment or intervention by leveraging a sharp discontinuity in the relationship between a continuous assignment variable and an outcome variable. In our context, the core concept of RD is based on the notion that the time-dependent indoor activity seasonality is inherently similar in the 8 weeks study period, except for the exposure to the wildfire event. By comparing the indoor seasonal activity for the affected counties at the starting date of the event, we can isolate the anomalous effect caused by wildfires from confounding factors (i.e., other potential local anomalies in the mobility).

For regression discontinuity analysis, we use a local linear regression model. It estimates the event’s effect by fitting a linear regression line separately for a set of observations after and before it. The local linear regression model can be represented as follows:

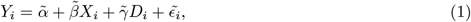

where:

- *Y*_*i*_ is the outcome variable for observation i;
- *X*_*i*_ is the assignment variable for observation i;
- *D*_*i*_ is the wildfire event indicator variable, such that:

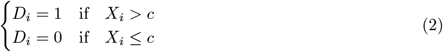

where *c* is the moment of impact of the event;

- 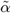 and 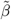 are the coefficients representing the intercept and slope of the regression line, respectively;
- 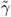 is the wildfire event effect, which represents the difference in outcome between the observations before and after the event;
- 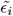 is the error term.

To give a larger weight to observations closer to the moment of the event, a weighting function is often applied in regression discontinuity analysis. One commonly used weighting function is the triangular kernel. The triangular kernel weighting function can be defined as:

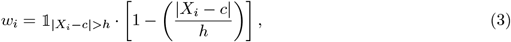

where:

- *w*_*i*_ is the weight assigned to observation *i*;
- *X*_*i*_ is the assignment variable for individual *i*;
- *c* is the moment of impact of the event;
- *h* is the bandwidth parameter that determines the width of the window around the moment of impact.

The weight *w*_*i*_ decreases as the distance between *X*_*i*_ and *c* increases, with observations exactly at the cutoff receiving the highest weight (*w*_*i*_ = 1) and observations further away receiving lower weights.

### 2.3 Infectious disease model

In our study, we develop a simple SIR model of disease spread to model infectious disease dynamics of the form:

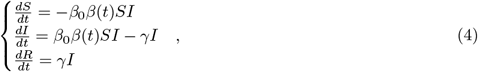

where:

- *S* represents the number of susceptible individuals;
- *I* is the number of infectious individuals;
- *R* is the number of recovered individuals;
- *γ* is the recovery rate;
- *β*_0_*β*(*t*) is the transmissibility rate, defined as the product of homogeneous time and country-independent parts *β*_0_ and *β*(*t*). *β*_0_ is a constant parameter that takes into account the overall transmissibility, while *β*(*t*) is related to the seasonality effect and the potential disruption of human mobility *σ*_*it*_ caused by wildfires.

We define the *force-of-infection* as 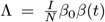 and the total population *N*. We run all the simulations starting one week before the start of the wildfire. In our study, the SIR model is evaluated for the *affected* counties OR, WA, and CA and the *unaffected* counties described in subsection 2.1. To compare the model on the two types of counties, we introduce the *relative Attack Rate AR*(*t*) as the relative cumulative number of infectious individuals at time *t* over the population size, and the *relative Peak Incidence PI*(*t*) as the relative variation of the occurrence of new cases of disease at the incidence peak, where the *relative* quantities are obtained wrt every element of the control-group counties and then statistically analyzed.

We explore the relative attack rate and the relative peak incidence comparing *affected* and *unaffected* counties using baseline *reproduction ratios* (*R* = *β/γ*) values of 1.3, 1.5 and 3. We choose such values because 1.3 and 1.5 are compatible with a seasonal spread of respiratory viruses such as influenza and currently SARS-CoV-2, and 3.0 was close to the basic reproduction ratio of the wild-type of SARS-CoV-2.

## 3 Results

### 3.1 Disruptions to mobility seasonality due to wildfires response

Given the disruptive smoke spread on U.S. West Coast in September 2020 (Figure 1, left-hand side), our preliminary analysis compares the AQI for *affected* counties in Oregon (OR), Washington (WA) and California (CA) (Figure 1, right-hand side). To provide a baseline, we report the AQI of U.S. *unaffected* counties. We notice that a peak in AQI is visible for OR, WA, and CA affected counties right after 2020/09/10. Oregon (OR) shows the highest peak.

**Figure 1:**
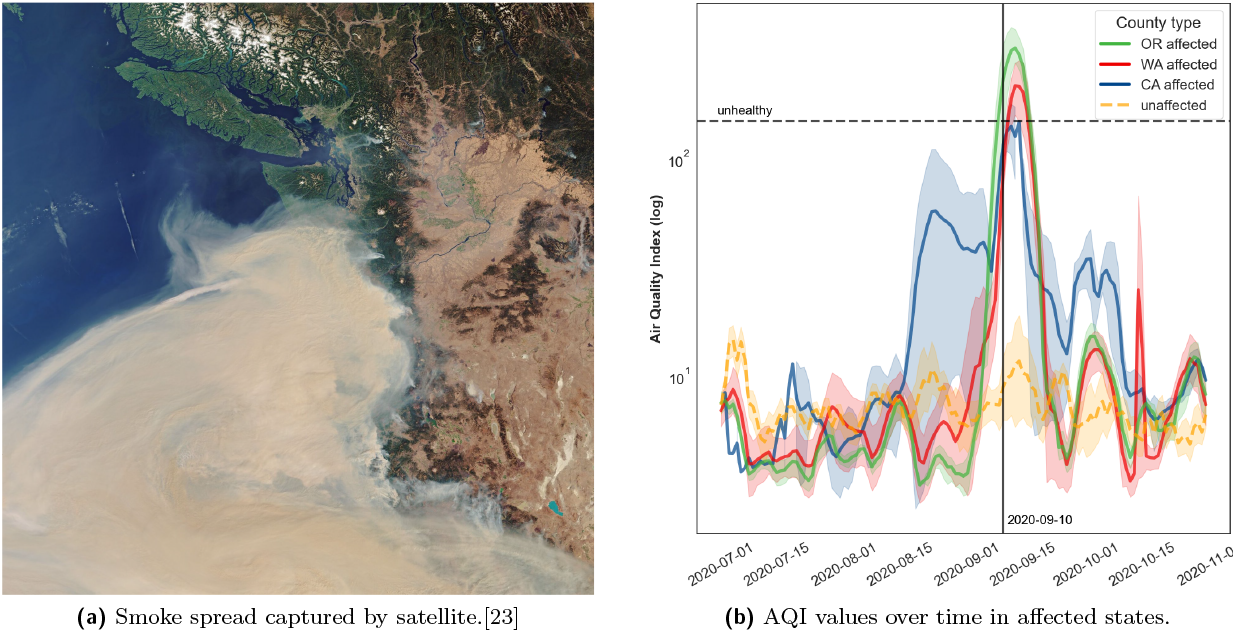
(a) Image captured on 2020/09/10, by Copernicus Sentinel-3, showing the extent of the smoke plume. (b) AQI for Oregon (OR), Washington (WA) and California (CA) counties affected by unhealthy air quality during the September 2020 wildfires. The reported AQI takes into account a 5-days moving average.

By exploring the variation of the indoor activity in the counties *affected* by the smoke of the September 2020 wildfires (Figure 2), we detect significant increases in the time spent indoors, compared to the baseline. These significant peaks are found in all the Oregon counties under study and in King, Clark, and Spokane countries in Washington. A slight signal of a peak in the indoor activity seasonality can also be noticed in the affected Plumas County, CA, even though its AQI is lower compared to that of the other *affected* counties considered. It can be noticed that, even when the peak of indoor activity is not significant with respect to the baseline in Washington counties, there is evidence of an increase in indoor activity.

**Figure 2:**
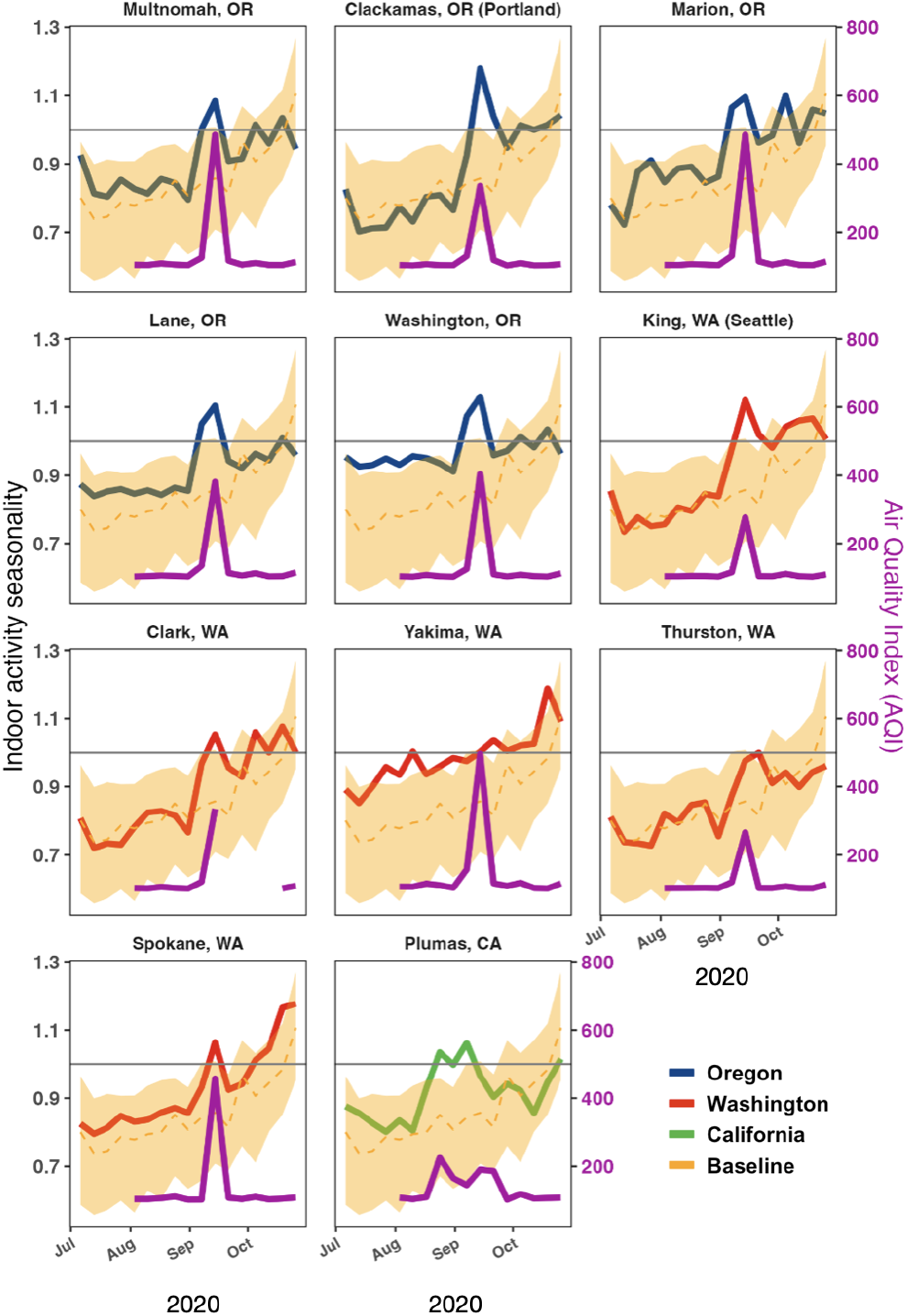
Indoor activity seasonality index between 2020/07/01 and 2020/11/01 in the 11 selected *affected* counties. In each subplot, we also take into account the median and 95% CI of indoor activity seasonality index for the baseline *unaffected* counties (yellow curves). Violet curves represent the AQI of the *affected* counties in the studied time period.

We quantified discontinuities in indoor activity seasonality with respect to September 2020 wildfires. Table 1 shows the results of the analysis. We observe that all Washington *affected* counties show a significant increase in indoor activity seasonality after the starting date of the wildfires. This also holds for Multnomah and Clackamas counties in Oregon. This latter is the county showing the greatest increase in indoor activity overall.

**Table 1:** Difference in 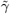 before and after 10 September 2020 and 90% CI.

| Country | County | $\Delta\tilde{\gamma}$ | $\Delta\tilde{\gamma}_{5\%}$ | $\Delta\tilde{\gamma}_{95\%}$ |
| --- | --- | --- | --- | --- |
| OR | Multnomah | 0.09 | 0.00 | 0.19 |
| OR | Washington | 0.07 | -0.01 | 0.14 |
| OR | Clackamas | 0.30 | 0.23 | 0.36 |
| OR | Lane | 0.07 | -0.01 | 0.14 |
| OR | Marion | 0.02 | -0.06 | 0.10 |
| WA | King | 0.15 | 0.10 | 0.20 |
| WA | Spokane | 0.13 | 0.08 | 0.18 |
| WA | Clark | 0.10 | 0.02 | 0.18 |
| WA | Thurston | 0.15 | 0.09 | 0.21 |
| WA | Yakima | 0.02 | 0.01 | 0.04 |
| CA | Plumas | 0.02 | -0.01 | 0.04 |

The increase is not significant for Washington, Lane, and Marion counties in Oregon and for Plumas County in California.

### 3.2 Impact on respiratory disease circulation caused by disruption on indoor activity

Figure 3 shows the variation of relative attack rate and relative peak incidence between the different *affected* counties and the *unaffected*. The results show relative values above zero for both epidemic outcomes, meaning that the attack rate and the peak incidence of *affected* counties are higher than those of *unaffected* counties. Washington, OR, and Yakima, WA have the highest relative values. On the contrary, Plumas, CA, presents negative relative values, as we would have expected from the different behavior of its corresponding indoor activity seasonality. *reproduction ratio* equal to 3.0 generally results in the highest relative attack rate, while the *reproduction ratio* equal to 1.3 leads overall to the highest relative peak incidence values.

**Figure 3:**
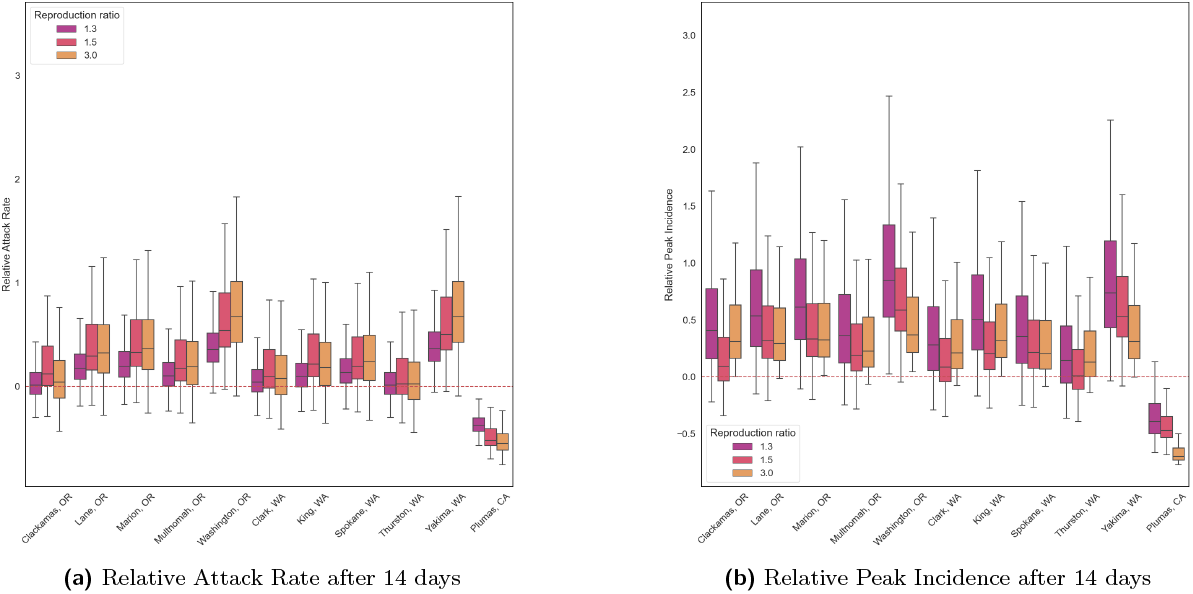
Relative attack rate (left) and relative peak incidence (right) for *affected* and *unaffected* counties after 14 days from the air quality alert due to wildfires (2020/09/10). We set the generation time equal to 14 days.

To characterize the impact of wildfires on different airborne respiratory diseases, we study model epidemic outcomes varying the generation time. We explore generation time ranging from 2 days (Flu-like) to 25 days (Pertussis-like) [32]. For this analysis, we focus on Yakima, WA (Figure 4). We study the variation in the relative values of attack rate and peak incidence compared again with the *unaffected* counties, while exploring different lengths of the generation time and three different *reproductive ratio* values. On the top row of the figure (*reproductive ratio = 1*.*3*), we observe a decreasing trend of the relative values of the epidemic’s outcomes. For short generation times (less than one week), we observe higher differences and also higher variability. We observe the same behavior, but with lower values of the relative measures, when we increase the *reproductive ratio* up to 1.5. This suggests that within generation times shorter than the period where the mobility of human activity occurs, the outbreak of the epidemic in an *affected* county is faster than in *unaffected* counties. The fact that within short ranges of generation time, the infections occur faster explains why we observe a higher variability of the results. When we look at higher values of the *reproductive ratio* (bottom row, Figure 4) we no longer see the same pattern. Since the epidemic grows faster, the disruption of human activity’s effect on the outbreak becomes very smooth.

**Figure 4:**
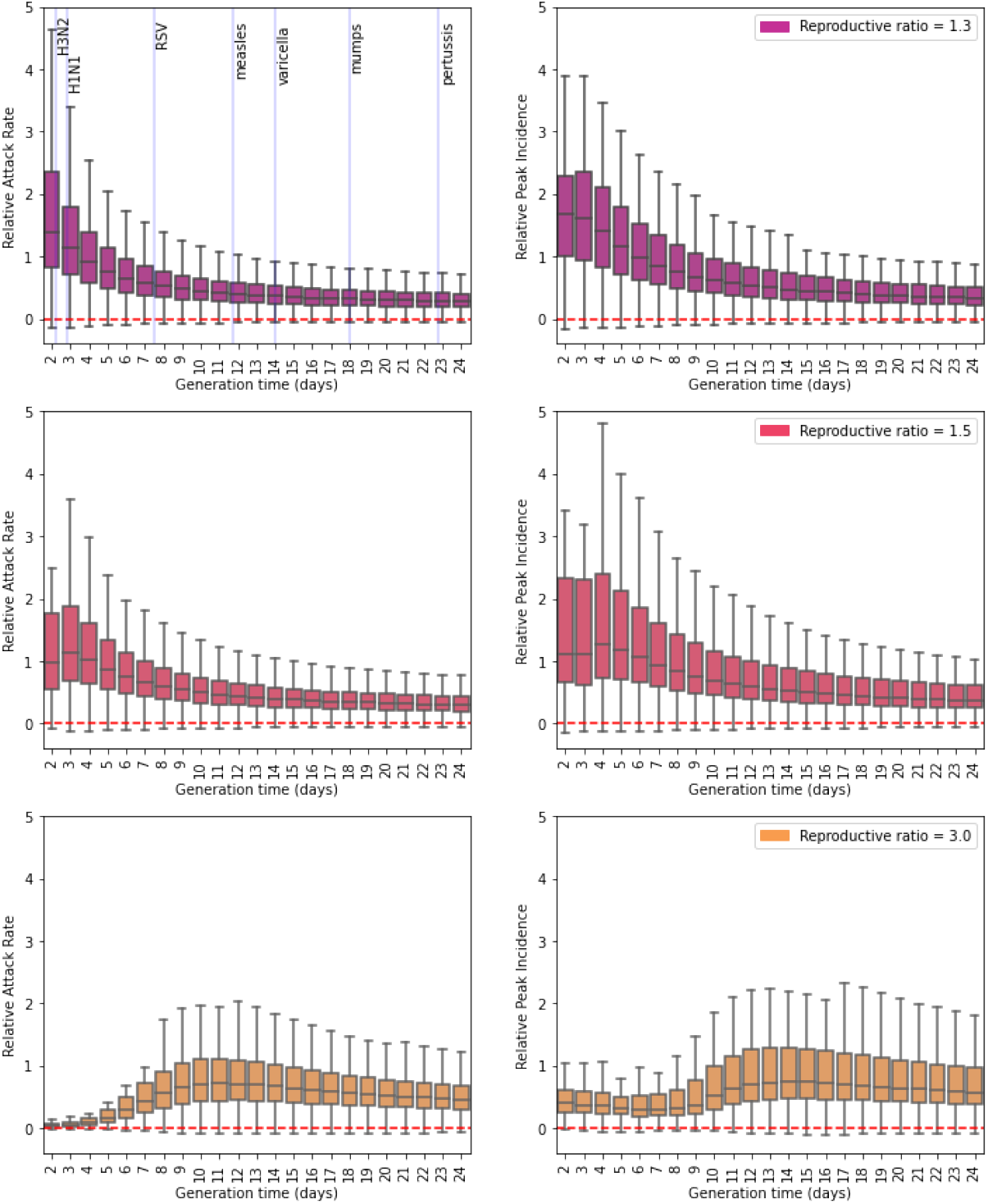
Relative attack rate (left) and relative peak incidence (right) after 14 days at one of the *affected* counties (Yakima, WA). On the top left panel, we show as a reference the mean serial interval for different airborne diseases [33].

## 4 Discussion

The impact of wildfires on public health extends beyond injuries. Smoke from wildfires can be transported by atmospheric circulation to areas unaffected by the fires, compromising air quality over vast stretches of land [3] and aggravating chronic respiratory conditions, like chronic obstructive pulmonary disease, emphysema, asthma [4]. But wildfire smoke may aggravate the burden of communicable respiratory disease, too. The perceived decline in air quality and adherence to safety guidelines can alter individuals’ ability and willingness to participate in outdoor activities, disrupting established behavioral patterns [18]. The disruption of human activity and the resulting influence on the transmission of communicable diseases is still unclear.

In our study, we aimed to bridge this gap by examining the effects of the severe wildfires that struck the West Coast of the United States of America in September 2020, and specifically the states of Oregon, Washington and California. Our focus was on assessing the impact of compromised air quality on human behavior and the potential consequences of the circulation of respiratory pathogens. We observed a significant increase in the time spent indoors in the affected areas of Oregon and Washington. This was instead not evident in California: we propose that this discrepancy comes from the fact that California residents are accustomed to regular exposure to wildfire smoke and Air Quality Index alerts, causing fatigue in their behavioral response. Then we studied the impact on epidemic circulation using a Susceptible-Infected-Recovered compartmental model to simulate the circulation of a respiratory pathogen right before, during and in the aftermath of the Air Quality Index alert, signaling compromised air quality. Specifically, we simulated the impact on the cumulative incidence of cases at four weeks after the event – called *relative attack rate* –, and on the peak daily incidence of cases over the same period – called *relative peak incidence*. We find that both the cumulative incidence and the peak daily incidence in *affected* counties are overall higher than those of *unaffected* counties in Oregon and Washington, not in California, which is consistent with the fact that no significant disruption in the time spent outdoors was visible there. In terms of *reproduction ratio*, a value of 3.0 (compatible with the SARS-CoV-2 onset) generally results in the highest relative cumulative incidence while a value of 1.3 (compatible with seasonal spreads of respiratory diseases) leads to the highest relative peak daily incidence, given a generation time of 14 days. Then we focused on a single *affected* county – Yakima Co., WA – and tested the impact of compromised air quality on disease circulation at varying average generation times. While with lower values of *reproductive ratio* we observed that, within short generation times, there is a higher difference of cumulative incidence and daily peak incidence between *affected* and *unaffected* counties, with higher values of the ratio the disruption of human activity’s effect on the outbreak becomes very smooth.

This study is preliminary and has limitations. We used a deterministic model of disease spread: this overlooks the inherent stochasticity of both natural hazard events and epidemic dynamics. Additionally, we study only one major wildfire event, and our findings may not be generalizable to similar events. Moreover, the study relies solely on the ratio of indoor-outdoor activities and does not incorporate information about disruptions in the time spent at home, which could affect epidemic circulation, too.

Our study is preliminary and educational in purpose. Notwithstanding, it highlights the need for additional modeling research on the impact that short periods of compromised air quality due to wildfire smoke may have on the circulation of respiratory pathogens. This is crucial to inform targeted interventions and awareness campaigns during wildfire seasons, emphasizing the importance of protecting vulnerable populations and promoting preventive measures. In particular, the proposed analysis underscores the importance of enhancing emergency preparedness protocols in regions prone to wildfires along with policy guidelines preventing epidemics spread.

## Data Availability

All data produced in the present study are available online and in the manuscript is cited the website in which to download it

https://github.com/bansallab/indoor_outdoor

https://www.epa.gov/outdoor-air-quality-data

## Acknowledgments

This work is the output of the Complexity72h workshop^3^, held at IFISC in Palma, Spain, 26-30 June 2023. We want to acknowledge Shweta Bansal for insightful discussions on the design of the project and for data provision.

## Footnotes

[1] All census data about the U.S. can be found in https://data.census.gov/

[2] https://github.com/bansallab/indoor_outdoor

[3] https://www.complexity72h.com

